# Performance of a Type 1 Diabetes Genetic Risk Score in a Multi-centric Study from India and its Implications in Clinical Practice

**DOI:** 10.64898/2026.02.21.26346764

**Authors:** Alagu Sankareswaran, Dimple Lavanuru, Bhanu Teja Nalluri, Shalbha Tiwari, Rashmi Nagaraj, Nagendra Khadri, Akila Prashant, Srinivas G N S V Kandula, Vedavati Purandare, Vaibhavishree Muniswamy, Navyashree Mugur Jagadeesha, Prajwal Guruswamy, Neelaveni Kudugunti, MR Savitha, Radhe Shyam Tapadia, Basavanagowdappa Hathur, Rakesh Kumar Sahay, Ambika G Unnikrishnan, Suraj S Nongmaithem, Bipin Sethi, Giriraj R Chandak

**Affiliations:** Genomic Research on Complex Diseases Group (GRC-Group), CSIR-Centre for Cellular and Molecular Biology (CSIR-CCMB), Hyderabad, India; Academy of Scientific and Innovative Research (AcSIR), Ghaziabad, India; Department of Endocrinology, Osmania Medical College, Hyderabad, India; Chellaram Diabetes Institute, Pune, India; Department of Paediatrics, JSS Medical College, JSS Academy of Higher Education & Research, Mysuru, India; Department of Paediatrics, Mysore Medial College and Research Institute, Mysuru, India; Department of Biochemistry, JSS Medical College, JSS Academy of Higher Education & Research, Mysuru, India; Department of Endocrinology, Tapadia Diagnostic Research Centre, Hyderabad, India; JSS Academy of Higher Education & Research, Mysuru, India; Lightening Lives LLP, Atal Incubation Centre, Hyderabad. India

**Author notes:** Senior authors (Basavanagowdappa Hathur, Rakesh Kumar Sahay, Ambika G Unnikrishnan, Suraj S Nongmaithem, Bipin Sethi, Giriraj R Chandak).

## Abstract

**Background:** Genetic risk scores (GRS) for type 1 diabetes (T1D) have been developed primarily in European populations, limiting their generalisability across ancestries. Indians differ from Europeans in clinical characteristics of T1D and overall genetic architecture, yet systematic evaluation of T1D GRS performance in multi-regional Indian cohorts is lacking.

**Methods:** The study included 597 T1D patients and 3347 non-diabetic controls from different regions in India. Genotyping, imputation, quality control analysis, and construction of the 67-SNPs T1D GRS were performed using standardised pipelines. Discriminative performance was assessed using Receiver Operative Curve-Area under Curve (ROC-AUC) analysis, and optimal thresholds were derived using Youden’s index. *HLA-DQ* diplotype frequencies were compared, and association analysis was conducted using multivariable logistic regression.

**Findings:** T1D GRS showed consistent discriminative performance across Indian cohorts [ROC-AUC=0.84 (range=0·78-0·87)], supporting its comprehensive use for T1D classification in India. Notably, its performance was lower in islet cell autoantibody (IA) negative compared with IA positive T1D patients (ROC-AUC, 0·75 vs 0·85) and in adult-onset than in childhood-onset patients (0·74 vs 0·84). We observed a lower frequency of protective *HLA-DQ* diplotypes and a strong association of HLA-DQ81 containing diplotypes in childhood-onset T1D. Application of an India-specific T1D GRS score improved the sensitivity than the European cut-off.

**Interpretation:** T1D GRS is a valuable unified diagnostic tool in Indians, but its performance varies by islet cell autoantibody status and age at onset, likely reflecting population-specific *HLA* architecture. European-derived T1D GRS thresholds under-classify the genetic risk, highlighting the importance of ancestry-aware optimisation in Indians.

**Funding:** CDRC grant CDRC202111026 and CSIR Intramural Grant P50.

**Research in context:** *Evidence before this study:* Previous studies have shown that a 67-SNPs T1D genetic risk score (GRS) can distinguish T1D patients from non-diabetic controls and other forms of diabetes, but its performance varies across ancestries. Islet cell autoantibodies (IA) have important diagnostic value for classifying type 1 diabetes (T1D). However, their prevalence in India varies widely, with up to one-quarter of patients testing negative, limiting their clinical utility. Evidence supporting the use of the T1D GRS in India, combined with IA antibodies status is limited to a single cohort representing one linguistic group. The applicability of T1D GRS across multi-centric clinical settings has not been systematically evaluated.

*Added value of this study:* This study validates the 67-SNPs T1D GRS across multiple Indian cohorts representing major linguistic groups, supporting its use as a unified diagnostic tool. Differences in T1DGRS performance between childhood-and adult-onset T1D are linked to enrichment of protective *HLA-DQ* diplotypes in adult-onset disease, providing genetic insight into disease heterogeneity. The study also demonstrates that European-derived GRS thresholds systematically under-classify genetic risk in Indians and the population-specific threshold is essential.

*Implications of all the available evidence:* The European-derived T1D GRS can be applied across Indian clinical settings with consistent discriminative performance. However, its utility is influenced by islet cell autoantibody status and the age at onset of disease. Ancestry-aware threshold optimisation substantially improves diagnostic accuracy and is essential for equitable implementation of T1D GRS in Indians. Larger studies are needed to identify population-specific risk variants and further refine genetic tools for clinical diagnosis.

## Introduction

Type 1 Diabetes (T1D) is a chronic autoimmune disorder characterised by the immune-mediated destruction of pancreatic β-cells, leading to an absolute deficiency of endogenous insulin production(1). The presence of islet cell autoantibodies (IA) is a defining feature of T1D, which plays a crucial role in disease classification and diagnosis(2). However, both the number and type of IA present vary substantially across populations, and are closely linked to the age at disease onset (3–5). Childhood-onset T1D is typically associated with multiple IA positivity and rapid β-cell loss, whereas adult-onset disease often presents with fewer IA positivity and greater clinical heterogeneity(6,7).

Genetic susceptibility plays a major role in T1D pathogenesis, with an estimated heritability of ∼80%(8), most of which is attributable to the Human Leukocyte Antigen (*HLA*) region(9). The classical risk haplotypes, DR3-DQ25 and DR4-DQ81, remain crucial to disease prediction, and landmark initiatives have used these markers to select high-risk individuals to identify autoimmune triggers(10). Beyond *HLA*, advances in genome-wide association studies (GWAS) have identified over 75 *non-HLA* loci linked to T1D, paving the way for the development of genetic risk scores (GRS)(11). In simple terms, a GRS aggregates the cumulative effect of risk alleles across multiple loci into a single quantitative measure, offering a scalable and clinically relevant tool for identifying genetic susceptibility(12). Several studies across European, East Asian, African and multi-ethnic populations have demonstrated the clinical utility of T1D GRS in differentiating T1D from other forms of diabetes and non-diabetic individuals(13–15).

Despite these advances, data from South Asia remains scarce. Our earlier work validated a 67-SNPs T1D GRS, in an Indo-European cohort from Western India, where it successfully discriminated T1D from Type 2 Diabetes (T2D) and non-diabetic controls, although, with a lower ability compared to Europeans(16). However, this evidence came primarily from a single cohort, which necessitates investigating the T1D GRS applicability in a multi-regional setting. Moreover, recent evidence suggests that GRS distribution may vary by population, advocating population-specific risk thresholds(17). Therefore, it is crucial to first identify the applicability of the T1D GRS while accounting for the local ancestry-related differences, if any, before implementing these scores in clinical practice.

In this study, we assessed the discriminative ability of the 67-SNPs T1D GRS in a multi-centric Indian cohort encompassing the country’s two primary linguistic groups, Indo-Europeans and Dravidians, to evaluate its suitability as a unified metric.

## Methods

### Type 1 diabetes cohorts

T1D was diagnosed in individuals with classical symptoms of hyperglycemia and history of diabetic ketoacidosis or ketonuria at presentation(18). Based on the age at onset, individuals were classified as childhood-onset T1D (<18 years) or adult-onset T1D (≥18 years). All clinically diagnosed individuals were on long term follow-up and on insulin treatment from the tertiary care centres representing one of the four cross-sectional cohorts namely Chellaram Diabetes Institute (CDI; n = 102) from Pune, Western India, Tapadia Diagnostic Research Centre (TDRC; n = 132), and Osmania General Hospital (OSM; n = 207) from Hyderabad, Southern India and JSS Academy of Higher Education and Research (JSS; n = 188) from Mysore, Southern India.

### Control cohorts

The parents of the children recruited in two prospective birth cohorts: Mysore Parthenon Cohort (MPC; n = 1047) and Mumbai Maternal Nutrition Project (MMNP; n = 2300) were used as non-diabetic controls for the study. The MPC was established in Mysore, South India in 1997 to study the long-term effects of maternal glucose tolerance and nutrition on offspring cardio-metabolic and cognitive outcomes(19). The non-diabetic status of the parents of this cohort was confirmed, using fasting glucose (WHO 1999 criteria) at the follow-up of children at 5-7 years of age. The MMNP cohort is a randomized trial in Mumbai slums, Western India (established in 2006), assessing the impact of pre-and peri-conceptual micronutrient supplementation on offspring cardio-metabolic health(20). The non-diabetic status of the participants was confirmed at follow-up of the children at 6 years of age.

### Islet autoantibodies and C-peptide measurement

Three islet cell autoantibodies namely glutamic acid decarboxylase 65 (GAD65), islet antigen 2 (IA2), and zinc transporter 8 (ZnT8) were measured using commercial ELISA kits (RSR Limited, Cardiff, UK). Individuals were classified as IA positive based on the following cut-offs: GAD65 ≥5 IU/mL, IA2 ≥7.5 IU/mL, and ZnT8 ≥15 IU/mL. Random serum C-peptide concentrations were measured using chemiluminescence immunoassays: Abbott ARCHITECT (CDI cohort), Siemens ADVIA Centaur (OSM cohort), and Roche Elecsys C-Peptide II (JSS cohort) at the respective sample collection centres.

### Sample preparation, high throughput genotyping, quality control and principal component analysis

Genomic DNA was extracted from peripheral blood samples using the QIAamp Midi DNA isolation kit (Qiagen) according to the manufacturer’s protocol. For all T1D patients, Infinium Global Screening Array (GSA) v3·0MD chips were used to generate genotype data (Illumina Inc., USA). Genome Studio 2·0 was used to call the genotypes and quality control (QC) of SNP data was done with filters: sample call rate ≥95% and gentrain ≥0·5, cluster separation ≥0·4 and AA/AB/BB Tdev <0·06. More stringent QC filters, such as SNP call rate ≥95%, Hardy-Weinberg equilibrium p-value <10^−6^ and minor allele frequency ≥ 0·5% were applied using plinkv1.9. A total of 32 samples failed in the QC assessment and were excluded from further analysis.

Fathers in MPC (n = 511) and 696 mothers in MMNP were genotyped using GSAv1·0 chips. Remaining individuals in the MMNP cohort were genotyped using GSAv3·0MD chips. The overlapping SNPs between both GSA versions were merged with the genotype data of the T1D patients, prior to QC analysis. For MPC mothers, high-throughput genotyping was performed using the Illumina Infinium CoreExome-24 Kit and the data were processed separately but subjected to the same QC as described above.

We applied a minor allele frequency cut-off of >5% to individual cohorts and merged overlapping SNPs from different chips. The LD pruning and PCA of the combined data were done using plinkv1·9. The projection PCA was conducted using Eigensoft v6·1·3 with the reference population data from Basu et al. 2016(21).

### Imputation and generation of genetic risk scores

The QC analysis provided high confidence data on 313,677 SNPs that were imputed using the NIH TOPMED R3 reference panel. The QCed data on 237,389 SNPs in MPC mothers was imputed separately using the same panel. The *HLA* alleles for the *DQA1* and *DQB1* locus were imputed separately using HIBAG R package at two-field resolution. A platform specific multi-ethnic GSK-HLARES reference panel was used for this imputation. Phasing of the *HLA-DQ* haplotypes (*DQA1 ∼ DQB1*) was done using the BIGDAWG R package.

Methods for generation of GRS have been described previously(13). Briefly, the GRS includes interaction of the *HLA-DQ* haplotypes, and linear scoring using plink for Other *HLA* (excluding *HLA-DQ* combinations) and *Non HLA* loci. The SNP rs1281943 was absent in the MPC mothers, and its frequency was <0·1% in the other control cohorts. We noticed that details on 10 SNPs were not available in the imputed data with R2 > 0·3. Hence we used proxy SNPs that showed complete linkage disequilibrium with sentinel SNPs (r^2^ = 1; D’ = 1) in South Asian data, obtained from TOPLD. No tag SNPs for rs540653847 could be identified in South Asians, hence a proxy SNP (rs571331280; r2 = 0·93; D’ = 0·98) from Europeans was used.

## Statistical analysis

The discriminative ability of T1D GRSs was evaluated by calculating the area under the curve (AUC) of the Receiver Operator Characteristic (ROC) curve (ROC-AUC). The confidence interval and the comparison of correlated ROC curves were estimated using the DeLong method. The pROC R package was used to estimate ROC statistics. The Wilcoxon rank-sum test was used for comparison of GRSs and clinical parameters across cohorts. Youden’s index (J = sensitivity + specificity − 1) was used to identify the optimal threshold by balancing sensitivity and specificity. Multivariable logistic regression was performed to compare adult and childhood-onset T1D using HLA-DQ diplotypes, adjusted for sex. All statistical analyses were conducted using R v4·2·0.

## Results

### Clinical and demographic characteristics of the T1D cohorts

This multi-centric study included T1D patients and non-diabetic controls from Western and Southern India. The T1D patients, aged between 1 to 53 years (median [IQR], 11·0 [7·0 - 15·0]), showed a lower proportion of males (40·8%) compared to females (P = 2·3 × 10⁻⁵). This was especially observed in childhood-onset T1D (P = 2·9 × 10⁻⁶). About 10% were classified as adult-onset T1D, in which most belong to the CDI cohort (61·4%), with equal male-female proportions. Duration of diabetes ranged from 0 to 36 years (median [IQR], 4·0 [1·0 – 7·0]).

Islet cell autoantibody profiles varied by cohort, with GAD65 being the most common. It was detected in 92% of OSM patients but in only 50% of TDRC patients, whereas it was between 68 - 77% in the JSS and CDI patient cohorts. Overall, IA positivity aligned with the range of 60 - 98%, reported in previous studies from India(22,23). Random C-peptide concentrations ranged between 3·3 to 2217·6 pmol/L (median [IQR], 26·5 [2·2-59·4]) (Supplementary Table 1). Only 10 patients had C-peptide concentrations above 600 pmol/L. Incidentally, they were also IA negative, supporting the accuracy of clinical diagnosis and suggest low chances of misclassification.

### Population substructure and allele frequency comparison

In order to reduce the analysis bias due to population-specific genetic architecture, we examined genome-wide ancestry patterns and allele frequency variations in 67 SNPs incorporated in T1D GRS. Principal component analysis (PCA) of T1D patients and controls identified a single dominant cluster indicating shared genetic background. Controls showed slightly greater dispersion along the first two PCs, likely due to larger sample size, while T1D patients formed a tighter cluster. Independent PCA for T1D patients and controls revealed similar patterns. Notably, T1D patients-only PCA suggested a subtle cline corresponding to the established North to South Indian genetic gradient. Projection PCA using mainland Indian references confirmed overlap with the Ancestral North Indian cluster, aligning with known admixture patterns. Importantly, this genetic structure was concordant to the linguistic classification, with the study participants aligning with their respective status as Indo-European or Dravidian linguistic groups. (Supplementary Figure 1).

Comparison of 67 SNPs between MMNP (Indo-European) and MPC (Dravidian) cohorts showed high concordance, with 62 SNPs differing by less than 0.025 risk allele frequency (RAF) and only five SNPs (rs41295121, rs6476839, rs9981624, rs10492166, rs9388489; all from *Non HLA* region) differed relatively more (RAF <0·035; Supplementary Figure 2). Wright’s Fst statistics further indicated minimal genetic differentiation (Supplementary Table 2). In the absence of distinct ancestry clusters, and a consistent allele frequency across 67 SNPs (used in the T1D GRS) in individual control cohorts, we used a combined control cohort for association analysis.

### Assessment of T1D GRS performance in Indian T1D patients

The ability of the T1D GRS to discriminate T1D patients (from four centres) and the common control group ranged from an ROC-AUC of 0·78 to 0·87 (Figure 1a). The combined multi-centric analysis showed an ROC-AUC of 0·84, and overlapping 95% confidence intervals indicated no statistically significant difference in T1D GRS performance across the centres. The ROC-AUC was highly comparable to our previous study (ROC-AUC [95%CI], 0·84 [0·82 - 0·85] vs. 0.83 [0·81 - 0·86]) (Figure 1b). Mean GRS values for T1D patients and controls were consistent across cohorts, with no significant within-group differences (Wilcoxon P > 0·05; Supplementary Figure 3).

**Figure 1:**
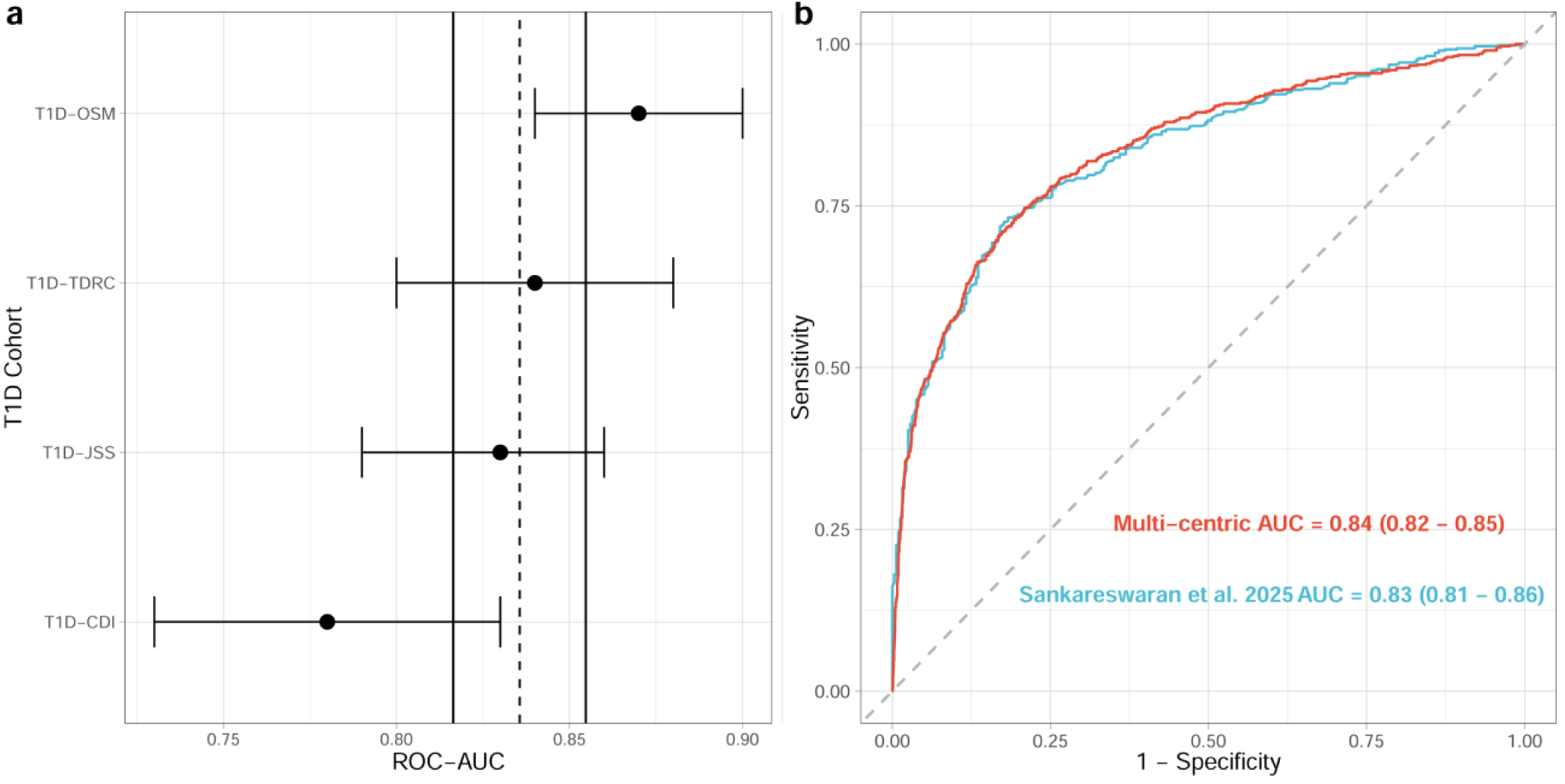
Discriminative ability of T1D GRS. **(a)** Forest plot showing the discriminative ability in individual T1D cohorts. The vertical dashed lines indicate the overall ROC-AUC, and the vertical solid lines indicate the corresponding confidence intervals of ROC-AUC. (**b)** ROC curve of the T1D GRS, from T1D and controls in the multi-centric cohort (red) and earlier single cohort study, Sankareswaran et al. 2025 (blue). T1D, Type 1 diabetes. GRS, Genetic risk score. AUC, Area under the receiver operating characteristic curve, along with 95% confidence intervals. ROC-AUC, Area under the receiver operating characteristic curve. CDI, Chellaram Diabetes Institute, TDRC, Tapadia Diagnostic Research Centre. JSS, JSS Academy of Higher Education and Research. OSM, Osmania General Hospital.

As reported previously, we observed that the T1D GRS performs better in IA positive patients than those negative for IA in distinguishing from controls (ROC-AUC 0·86 vs 0·75). Interestingly, the differences in the discriminative ability of T1D GRS was primarily driven by the *HLA* markers, and the Non HLA markers showed similar performance regardless of the antibody status (Supplementary Table 3).

### Differences in HLA-DQ haplotype frequencies among T1D cohorts

To understand the role of HLA variations, we compared the frequencies of 12 HLA-DQ *(DQA1∼DQB1)* haplotypes used in the T1D GRS with a frequency ≥0·5% in T1D cohorts or controls (Supplementary Figure 4; Supplementary Table 4). The well-known T1D risk haplotypes, DQA1*05:01∼DQB1*02:01 (DQ25; 40·2%) and DQA1*03:01∼DQB1*03:02 (DQ81; 23·8%), were most common in our T1D cohorts. The haplotype distribution was similar across T1D cohorts, aligning with prior findings, but subtle differences were evident. In the CDI cohort, DQ81 was about 10% lower, and DQ25 was less frequent (5·0% lower), while protective haplotypes like DQ61 were slightly more common (3·1% higher) than others cohorts. These differences may explain the reduced, though non-significant, discriminative ability of T1D GRS in the CDI cohort, possibly due to a higher proportion of adult-onset T1D cases (34·7% vs 4·7%; Supplementary Figure 5). Overall, major risk haplotypes were enriched across cohorts, but minor differences in the haplotype frequency are also important.

### Lower T1D GRS and distinct *HLA-DQ* diplotype patterns between childhood and adult-onset T1D

We evaluated the utility of the T1D GRS in both childhood-onset and adult-onset T1D patients within our cohorts. The mean GRS in adult-onset T1D patients group was significantly lower compared to childhood-onset T1D group (12·15 vs 13·25, P = 5·0 x 10^-4^, Figure 2a), leading to a lower discriminative ability in identifying adult-onset T1D patients (ROC-AUC, 0·74 vs 0·84, P = 3·9 x 10^-3^, Figure 2b). Consistent with this observation, analysis of *HLA-DQ* haplotype combinations (termed as diplotypes), revealed an enrichment of protective diplotypes in adult-onset compared with childhood-onset T1D patients (Supplementary Figure 6).

**Figure 2:**
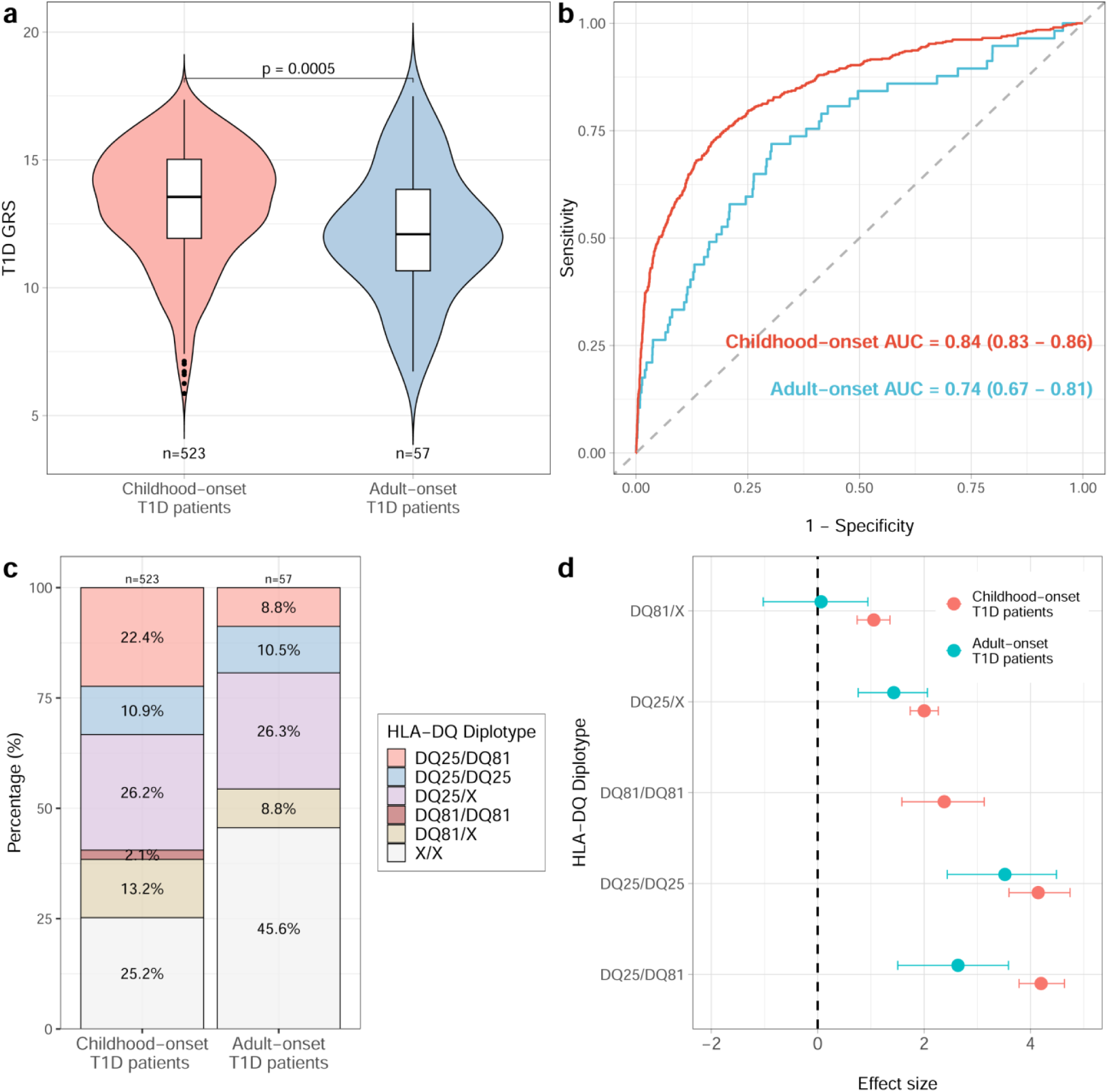
Comparison of the genetic factors between childhood and adult-onset T1D. **(a)** Violin plot comparing the T1D GRS between childhood and adult-onset T1D. **(b)** Discriminative ability of T1D GRS in childhood and adult-onset T1D**. (c)** Stacked bar plot of HLA-DQ diplotypes stratified by childhood and adult-onset T1D. X represents any haplotype other than DQ25 or DQ81. **(d**) Forest plot showing the association status of T1D-associated HLA-DQ risk diplotypes. T1D, Type 1 diabetes. GRS, Genetic risk score. AUC, Area under the receiver operating characteristic curve, along with 95% confidence intervals.

To further dissect the effects of specific *HLA* diplotypes, we focussed on DQ25 and DQ81, and X (for all remaining haplotypes) haplotypes. We observed a higher frequency of diplotypes containing DQ81 (DQ25/DQ81, DQ81/DQ81 and DQ81/X) in childhood-onset T1D patients than in adult-onset T1D patients. (Figure 2c; Supplementary Table 5). In contrast, the diplotypes containing at least one DQ25 without DQ81, (DQ25/DQ25 and DQ25/X) occurred at comparable frequencies in both groups. A sensitivity analysis excluding T1D patients who were negative for islet cell autoantibodies or had C-peptide concentrations missing or above 600 pmol/L showed similar patterns (Supplementary figure 7). Association analysis of T1D patients and controls indicated that homozygous diplotype of DQ25 (DQ25/DQ25) and the compound heterozygous DQ25/DQ81 confer the highest genetic risk for T1D in Indians. Additionally, the heterozygous diplotype containing exactly one DQ25 (DQ25/X) showed twice the effect than another heterozygous diplotype containing exactly one DQ81 (DQ81/X), confirming its widely reported role as one of the principal driver of T1D risk in Indians. Interestingly, DQ81 containing diplotypes but without DQ25, (DQ81/X and DQ81/DQ81) were independently associated only in childhood-onset T1D, despite having a lower overall risk compared to DQ25 diplotypes. Also, the heterozygous risk diplotype (DQ25/DQ81) has a significantly lower effect size in the association analyses of adult-onset T1D patients than in the childhood-onset T1D patients (Figure 2d, Supplementary table 6). Taken together, it may be surmised that while DQ25 is the predominant risk haplotype, DQ81 also play a significant role in determining early onset of the disease.

### T1D GRS thresholds for optimal discrimination in Indians

Having made some progress in identifying factors affecting the performance of T1D GRS in Indian T1D patients, we investigated the suitability of European T1D GRS centiles to Indian T1D patients. We noted a clear leftward shift of T1D GRS distribution in Indians. Only 30% (n = 175) of Indian T1D patients had T1D GRS above the European 50^th^ centile (T1D GRS = 14·60), compared with 50% expected under a matched distribution. Similarly, the European 90^th^ centile (T1D GRS = 16·54), a threshold, widely used to identify individuals at high genetic risk, was observed in only 4% (n = 23) of Indian T1D patients, substantially lower than the expected 10% (Supplementary Figure 8; Supplementary Table 7).

Following up from the above results and leveraging the availability of data from multiple Indian T1D cohorts, we derived the T1D GRS thresholds for Indians using Youden’s index. The maximum Youden’s index (J = 0·54) was observed at a T1D GRS of 11·85, correctly classifying 74·7% of clinically diagnosed Indian T1D patients (Figure 3a). In contrast, application of the established European cut off (T1D GRS = 13·00) correctly classified only 56·4% of Indian T1D patients. The European threshold demonstrated higher specificity (91·1%), as expected given its higher cut-off value. However, the corresponding Youden’s index was 0·47, which was significantly lower than that observed with the Indian-optimised threshold. Given the reduced discriminatory performance of the T1D GRS in IA negative and adult-onset T1D patients, we repeated the analyses in childhood-onset T1D patients, who were positive for islet cell autoantibodies (n = 411; Figure 3b). In this subgroup, the optimal cut off increased to 12·24, with an improved Youden’s index (J = 0·59). Even in this clinically stringent subgroup, classification using the Indian-specific threshold remained superior to the European cut-off (75·7% vs 61·8% sensitivity), strongly favouring use of India-specific T1D GRS cut-off for robust discrimination.

**Figure 3:**
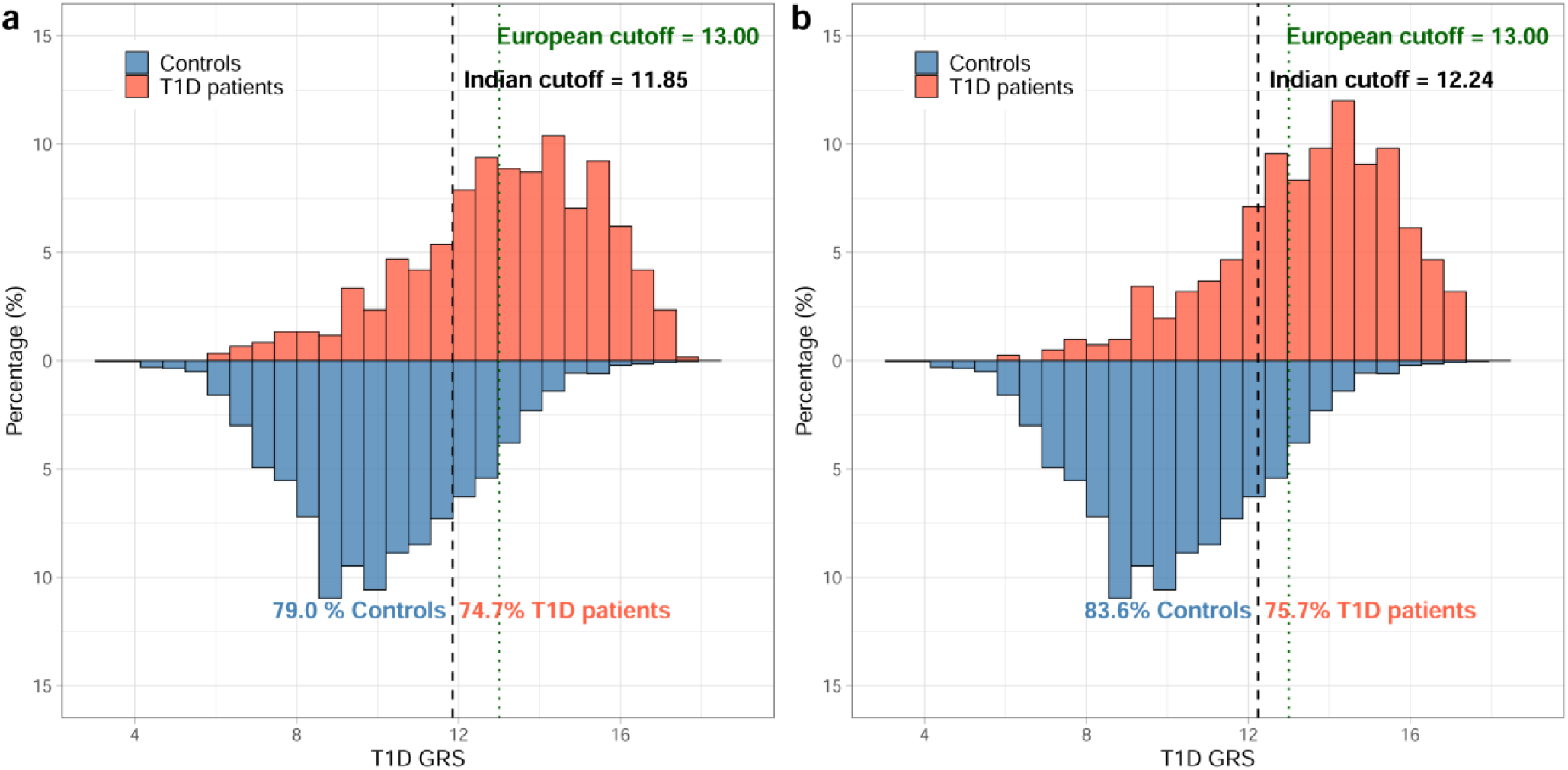
T1D GRS thresholds and optimal cut-off in Indian T1D patients. **(a)** Using all clinically diagnosed T1D patients and controls. **(b)** Using only islet autoantibody positive and childhood-onset T1D patients and controls. Black dashed line indicates the optimal cut-off point, determined using best Youden’s index from Indians. Green dotted line indicates the optimal cut-off from Europeans (taken from *Sharp et al 2019*). T1D, Type 1 diabetes. GRS, Genetic risk score.

## Discussion

The incidence of type 1 diabetes is rising globally and is projected to increase by 66% by the year 2040, with a disproportionate burden in low-income countries(24). With expansion of the clinical spectrum of T1D, the diagnostic challenges have intensified, particularly in resource-limited settings, underscoring the need for accessible tools to support an accurate diagnosis. In this context, T1D genetic risk score has emerged as a valuable marker for distinguishing T1D from other types of diabetes (13,25). Although its performance is lower in non-European populations, it remains a robust predictor of T1D across different ancestries(14).

Following on from our previous study demonstrating a lower discriminative ability of T1D GRS in Indians using a single cohort comprising Indo-European individuals(16), we conducted this multi-centric study including T1D patients and controls from multiple geographical regions representing Indo-European and Dravidian linguistic groups to explore the wider utility of T1D GRS. We highlight four main observations from this study: (A) The T1D GRS demonstrates lower but consistent discriminative ability across various centres; (B) it discriminates the IA positive T1D patients more effectively than the IA negative patients from controls; (C) distinct HLA-DQ haplotypes and diplotypes influence disease susceptibility and age at onset showing a greater role in childhood-onset T1D; and (D) the European-derived T1D GRS cut-offs underestimate T1D risk in Indians, and India-specific cut-offs identify T1D patients with better sensitivity.

Despite minor differences in ROC-AUC values, the discriminative ability of the T1D GRS across multiple cohorts representing two major linguistic groups was consistent. Although these cohorts do not capture the entire population diversity of India, they include urban referral centres from multiple regions and therefore provide a realistic view of genetic variability, particularly at T1D associated loci, and consequently the T1D GRS in Indians. The Indo-European and Dravidian groups are reported to exhibit distinct genetic, dietary, and microbiome profiles(26,27). However, the admixed population substructure of the cohorts and the absence of differences in risk allele frequencies across controls further support the applicability of the existing T1D GRS across India. Considering that the overall discriminative ability of the same T1D GRS in Indians (ROC-AUC 0.84) is lower than that of the Europeans (ROC-AUC 0·92), there is a need for larger studies from diverse parts of India to identify specific markers and improve its performance in Indians(13).

Islet cell autoantibodies have remained the strongest diagnostic marker for T1D since their discovery. However, recent studies report a growing proportion of IA negative T1D patients, posing a major diagnostic challenge in India, where this subtype is more frequent than in European cohorts(4). In this study, the T1D GRS retained meaningful discriminative ability in the IA negative group, which would be misclassified otherwise. A similar pattern has been reported in the African and US cohorts with varying proportions of IA negative T1D patients(5). Together, these findings suggest that the emerging non-autoimmune T1D subtype remains insufficiently studied. Large, well-powered studies are needed to identify its genetic and environmental determinants to support more inclusive diagnostic strategies.

Given the strong heritability of T1D driven by the *HLA* region, we examined the association of the *HLA-DQ* risk haplotypes used in the T1D GRS. Our findings reaffirm that DQ25 remains the principal risk haplotype in Indian T1D patients, showing higher effect size than the another risk haplotype, DQ81(28). However, on comparing children and adults, DQ81 emerged as a key risk determinant of childhood-onset T1D, which is consistent with literature linking DQ81 to rapid progression and multiple autoantibody positivity, a pattern more common in childhood-onset T1D (29). Notably, DQ81 is associated with all stages of T1D progression, whereas DQ25 appears restricted to progression from dysglycemia (stage 2) to symptomatic T1D (stage 3)(30), which may explain the dominance of DQ25 in Indians irrespective of onset age. Independent longitudinal studies with large sample sizes are required to explore age-specific gene-gene interactions and identify additional genetic markers.

Recent studies show that it is not appropriate to extrapolate the GRS developed in European populations to other ancestries without replication studies and careful adjustments(17). Using the T1D GRS as an example, we demonstrate the need for such an assessment, particularly as polygenic scores are increasingly used across diverse populations. The leftward shift in the distribution of T1D GRS observed in Indians in comparison to the Europeans underline the differences in effect sizes at known loci or the contribution of population-specific risk variants, not captured in the European studies. A similar shift to the left was noted in only IA positive childhood-onset T1D patients, suggesting that European-derived T1D GRS thresholds systematically under-classify genetic risk in Indians and highlight the substantial gains in ancestry-aware optimisation. Similar patterns have been reported in multi-ancestry controls, although their clinical implications have not been well established(17), as noted that the lower range of T1D GRS led to a lower cut-off for best discrimination in Indian T1D patients. Thus, European T1D GRS cut-off for discriminating T1D patients is fraught with the danger of missing a significant proportion of true T1D patients. A recent evidence from African population also advocates the usage of population-specific T1D GRS for best performance(31)

Further, we attribute the difference in the discriminative performance of T1D GRS in the CDI cohort compared to other cohorts as a reflection of the influence of variation in age at diagnosis rather than population-specific effects, as our earlier work from the same region demonstrated a good discriminative accuracy (16). The inverse relationship between T1D GRS and age of onset aligns with observations from the ADDRESS2 study(32). Conversely, the StartRight study reported no such differences, likely due to inclusion of late-onset adults only (median age 35 years)(33). These considerations are particularly relevant in India, where T2D often occurs at a younger age than in Europeans, complicating clinical differentiation of adult-onset T1D from T2D. Collectively, these evidences mandate a shift toward ancestry-aware GRS and polygenic scoring frameworks for equitable translation across global populations.

The major strength of this study is the inclusion of multiple cohorts from different geographical regions representing two major linguistic groups in India. Further, in the background of the admixed nature of individuals in all cohorts leaves no doubt about the utility of T1D GRS across the country. More replication studies from other geographical regions may strengthen this observation. One limitation of this study is the lack of replication of the association patterns between childhood and adult-onset T1D due to limited cohort sizes, but strong literature evidence supports the differential role for these haplotypes.

In conclusion, probably in one of the largest South Asian study, across multi-regional cohorts in India, we validate the utility of the European-derived T1D GRS in Indian populations. We also highlight important genetic differences linked to islet cell autoantibody status and the age at onset, and demonstrate the value of population-specific T1D GRS thresholds for clinical implementation. Together, these findings support a careful but forward looking integration of genetic tools into clinical decision making for diagnosis and management of type 1 diabetes.

## Other article information

### Contributors

GRC, SSN, AS and BS conceptualised the study; BS, SGK, AS, DL, BTN, RKS, RN, BH, AP, NK, AG, ST, and VP did data curation; AS, DL, VM, NMJ, PG conducted the formal analysis; DL, SSN, BS, RKS, AP, ST were responsible for the investigations presented within the study. AS, BS, SGK, BTN, RKS, RN, NK, AG, ST, VP and SSN, designed the methodology. GRC, SSN, BS, RKS, BH, AP, AG, ST did project administration and resource allocation. AS and SSN were responsible for the choice of software for appropriate analyses. GRC was responsible for overall supervision. AS wrote original draft of the manuscript. All authors critically reviewed and approved the final manuscript. GRC is the guarantor of the work and takes responsibility for data integrity and accuracy of the data analysis.

### Ethics declaration

The collection of clinical data and use of bio-banked samples for the biochemical, immunological, and genetic measurements was sanctioned by the Ethics committee of the respective collaborating institutes. CSIR-CCMB (IEC-92/2022), TDRC (IHEC-TDRC-HYD/1/2022), CDI (CDI/BR/2024/058), JSS (JSSMC/IEC/19052022/01NCT/2022-23), OSM (IEC-BHR/OMC/M.NO(07)/P-89).

### Statement on AI use

During revision of the manuscript, ChatGPT (OpenAI) was used solely for language editing and refinement. After using this tool, we reviewed and edited the content as required and take full responsibility for the content of the publication.

## Acknowledgements

We thank the participants in the study. The funding from Chellaram Diabetes and Research Centre (CDRC), Pune, India and the Council of Scientific and Industrial Research (CSIR), Ministry of Science and Technology, Government of India, New Delhi, India is gratefully acknowledged. GRC acknowledges support for JC Bose Fellowship awarded by Anusandhan National Research Foundation (ANRF), Department of Science and Technology, Government of India, New Delhi, India. We thank Prof. Caroline H D Fall from University of Southampton, Southampton, UK, Dr. Kalyanaraman Kumaran and Dr. G V Krishnaveni from Holdsworth Memorial Hospital, Mysuru, India and Dr. Sirazul A. Sahariah from Centre for the Study of Social Change, Mumbai, India for providing access to their cohorts and for their support in facilitating this work.

## Data Availability

Requests to data access should be submitted to the corresponding author Dr. Giriraj R. Chandak.

## Code Availability

All analyses used open source software such as R (version 4.2.0), pROC (version 1.17.0), ROCit (version 2.1.1), HIBAG (bioconductor version release 3.16), BIGDAWG (version 3.0.3) and PLINK (version 1.90b6.24)

## Supplementary figures

**Supplementary Figure 1:**
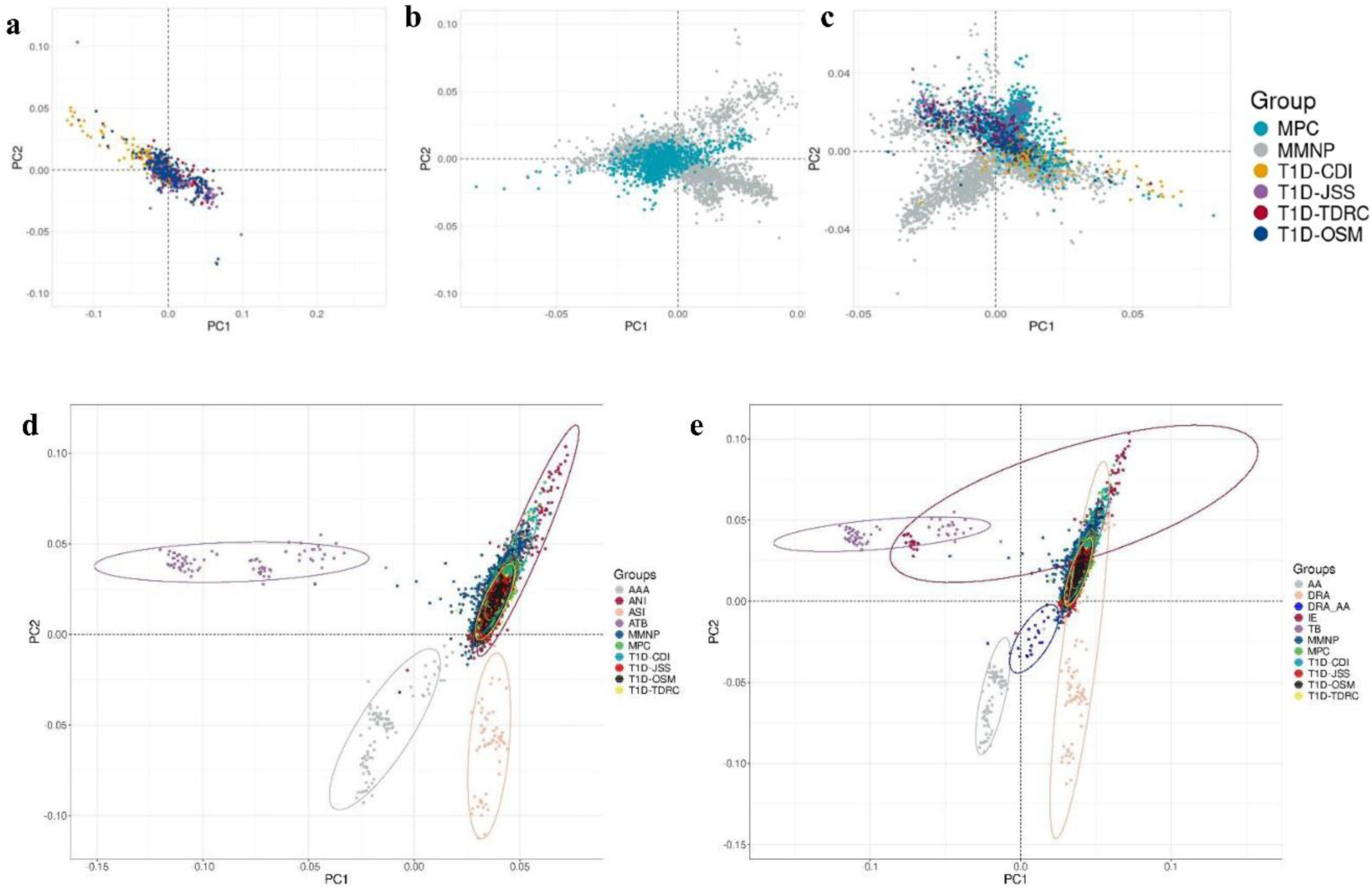
Scatter plots of the samples used in the study. **(a)** Principal component analysis (PCA) of only T1D cohorts **(b)** PCA of only control cohorts **(c)** PCA of all cohorts together. All three panels share the same color legend, as mentioned in c. **(d)** Projection PCA of the study data on Indian ancestral populations. AAA, Ancestral Austro Asiatic. ANI, Ancestral North Indian. ASI, Ancestral South Indian. ATB, Ancestral Tibeto Burman. **(e)** Projection PCA of the study data on Indian linguistic groups. AA, Austro Asiatic. DRA, Dravidian. DRA_AA, Dravidian austroasiatic, IE, Indo-European. TB, Tibeto Burman. T1D, Type 1 diabetes. CDI, Chellaram Diabetes Institute, TDRC, Tapadia Diagnostic Research Centre. JSS, JSS Academy of Higher Education and Research. OSM, Osmania General Hospital. MMNP, Mumbai Maternal Nutrition Project. MPC, Mysore Parthenon Cohort.

**Supplementary Figure 2:**
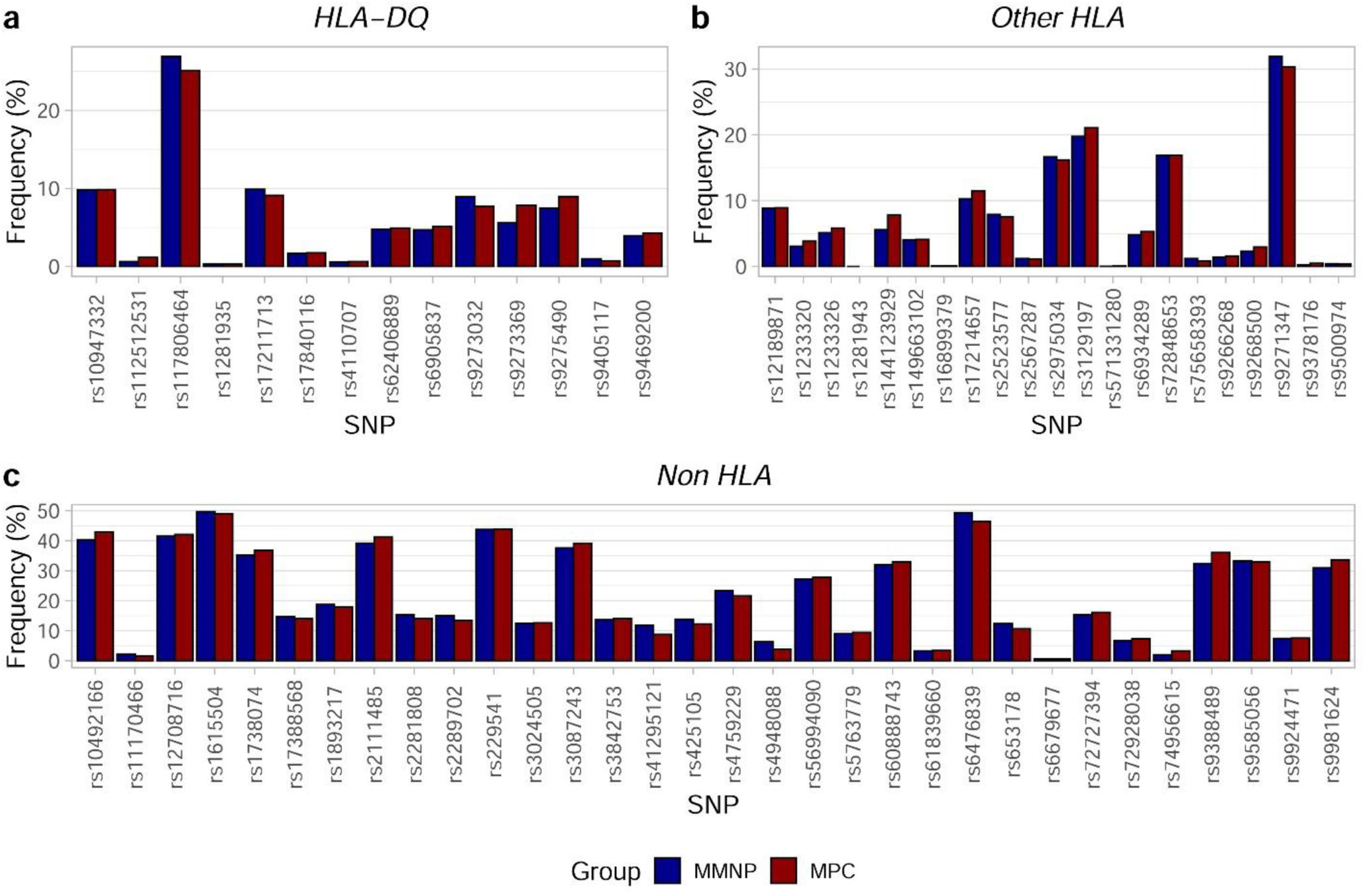
Comparison of the risk allele frequency of the 67 SNPs included in the T1D GRS among the control cohorts. (a) *HLA-DQ* SNPs (b) Other *HLA* SNPs C) Non *HLA* SNPs. T1D, Type 1 diabetes. GRS, Genetic risk score. *HLA,* Human leucocyte antigen. MMNP, Mumbai Maternal Nutrition Project. MPC, Mysore Parthenon Cohort.

**Supplementary Figure 3:**
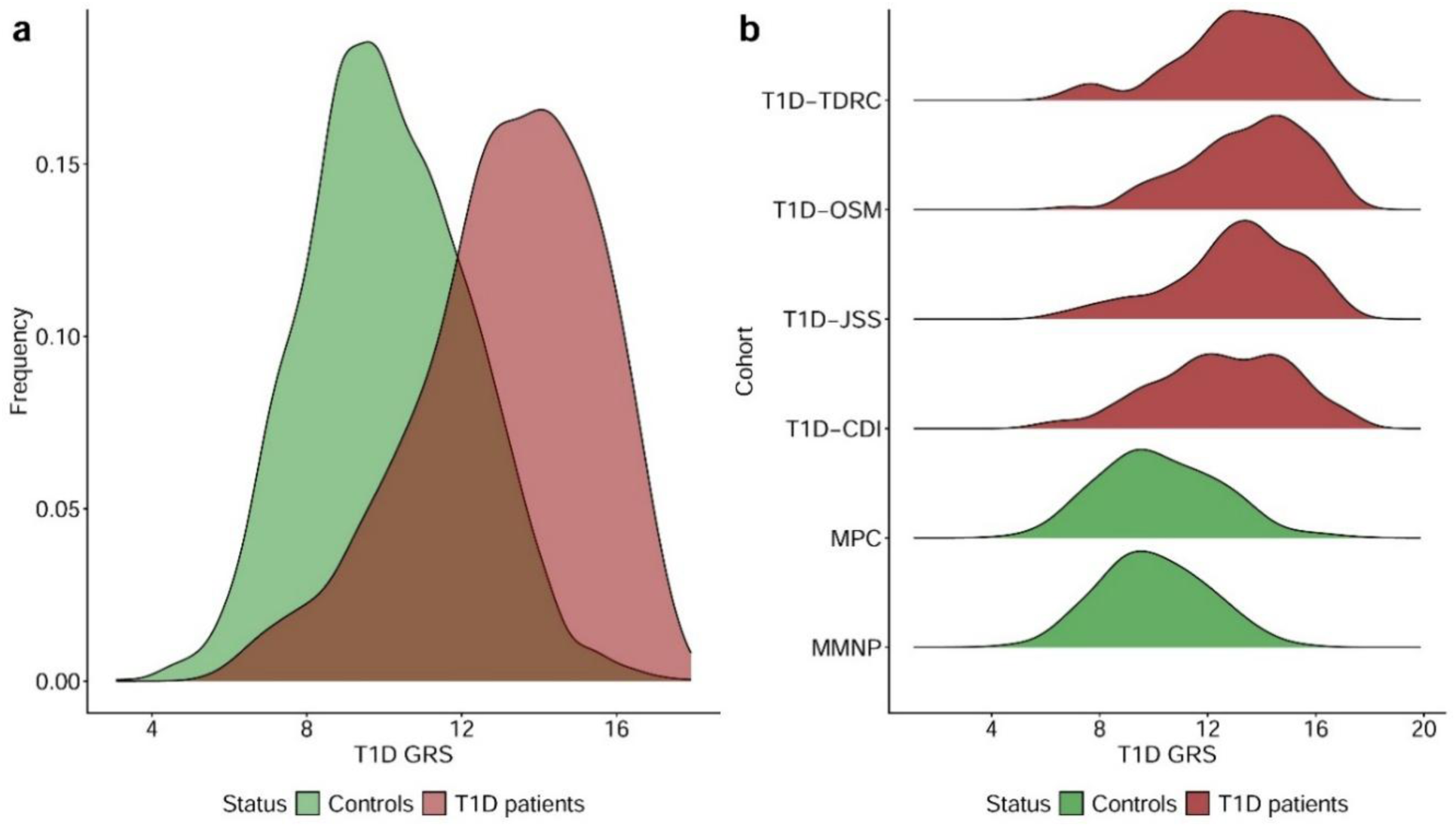
Distribution of T1D GRS in T1D patients and controls. **(a)**Density plot of T1D GRS across multi-centric T1D patients’ and control cohorts**. (b)** Ridge plot showing the distribution of individual cohorts. T1D, Type 1 diabetes. GRS, Genetic risk score. CDI, Chellaram Diabetes Institute, TDRC, Tapadia Diagnostic Research Centre. JSS, JSS Academy of Higher Education and Research. OSM, Osmania General Hospital. MMNP, Mumbai Maternal Nutrition Project. MPC, Mysore Parthenon Cohort.

**Supplementary Figure 4:**
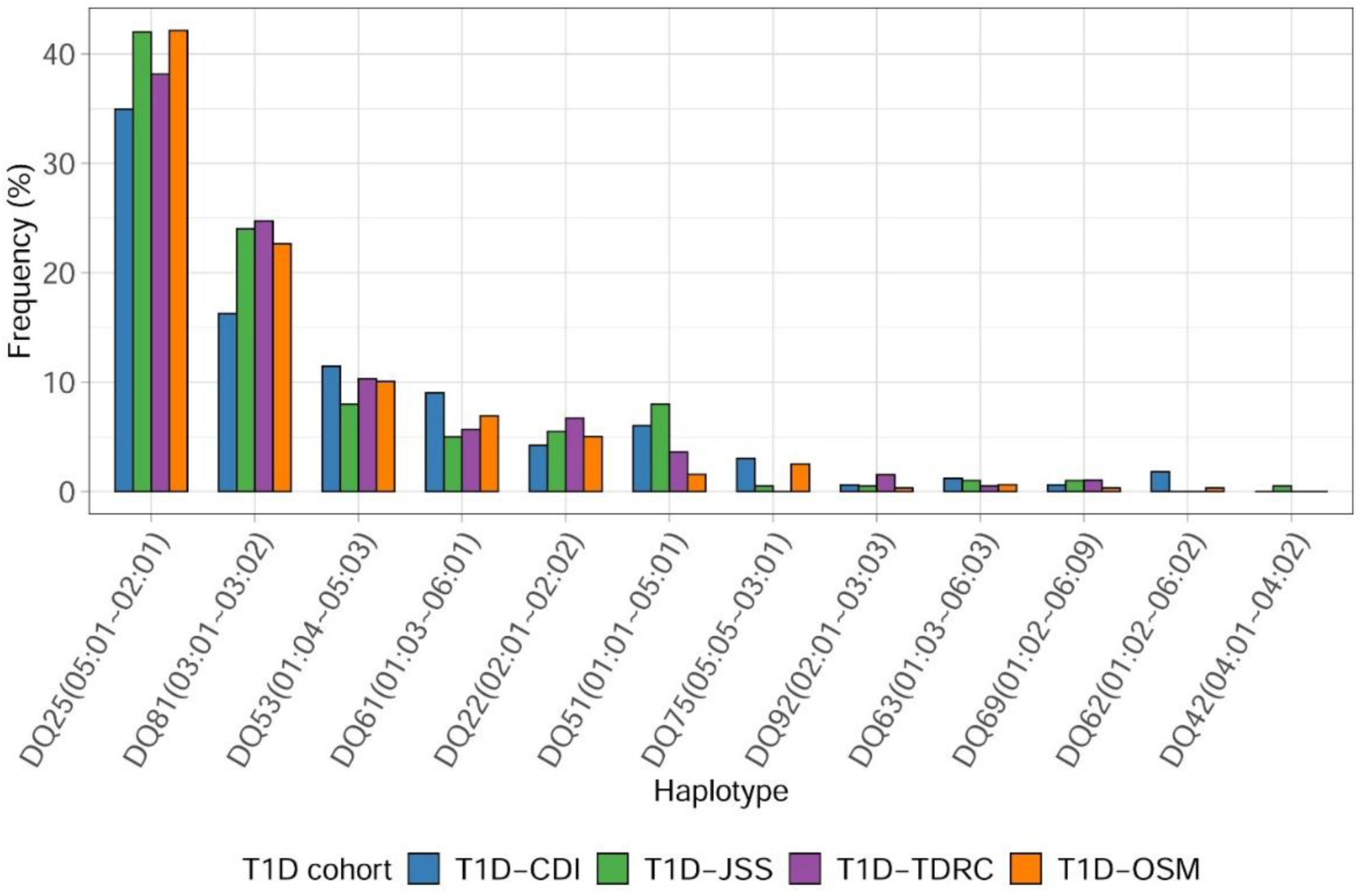
Frequency distribution of the *HLA-DQ* haplotypes used in T1D GRS in individual T1D cohorts. T1D, Type 1 diabetes. CDI, Chellaram Diabetes Institute, TDRC, Tapadia Diagnostic Research Centre. JSS, JSS Academy of Higher Education and Research. OSM, Osmania General Hospital.

**Supplementary Figure 5:**
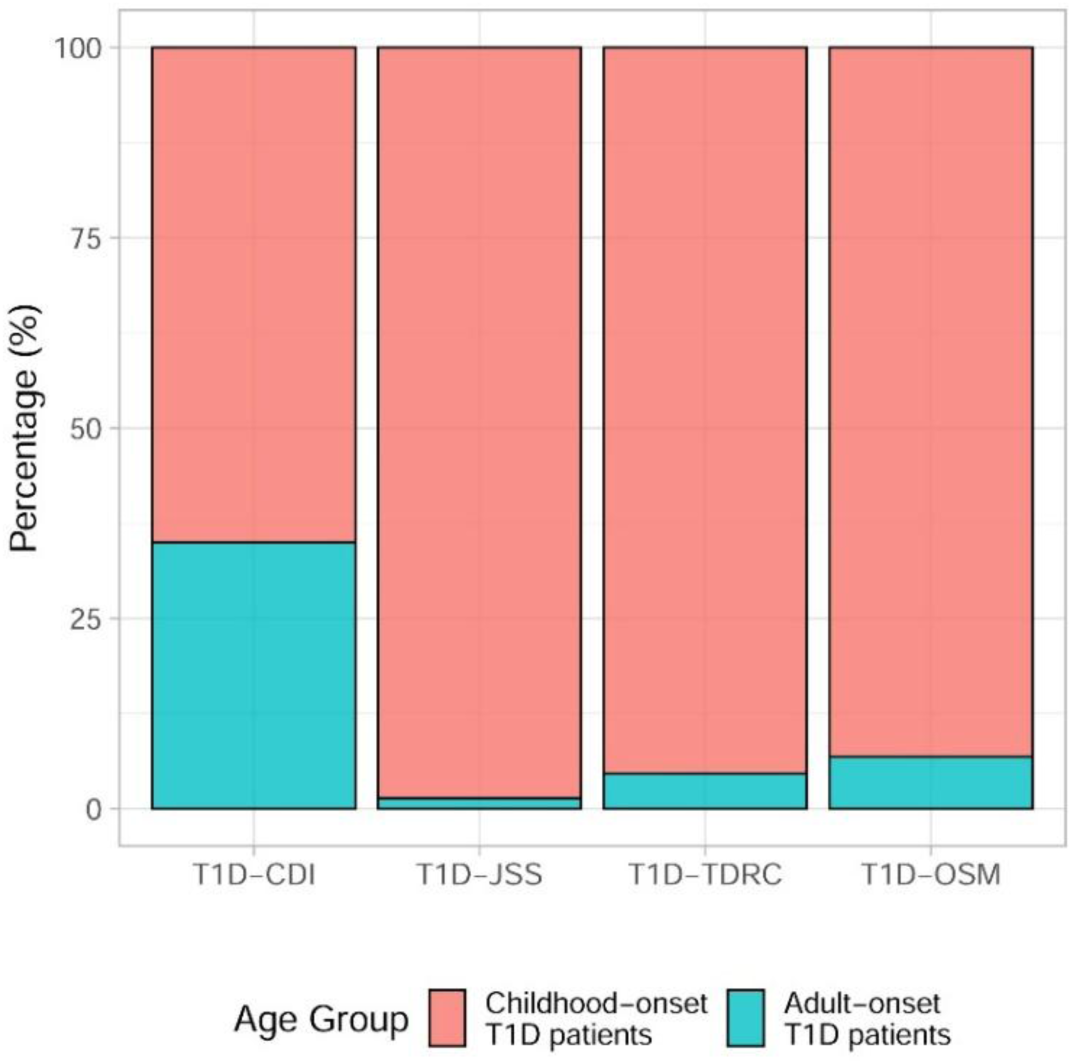
Comparative bar graph of age at diagnosis amongst type 1 diabetes (T1D) cohorts. Childhood-onset and adult-onset T1D patients are coloured in red and blue respectively. CDI, Chellaram Diabetes Institute. TDRC, Tapadia Diagnostic Research Centre. JSS, JSS Academy of Higher Education and Research. OSM, Osmania General Hospital. Age at diagnosis is missing in 17 T1D patients.

**Supplementary Figure 6:**
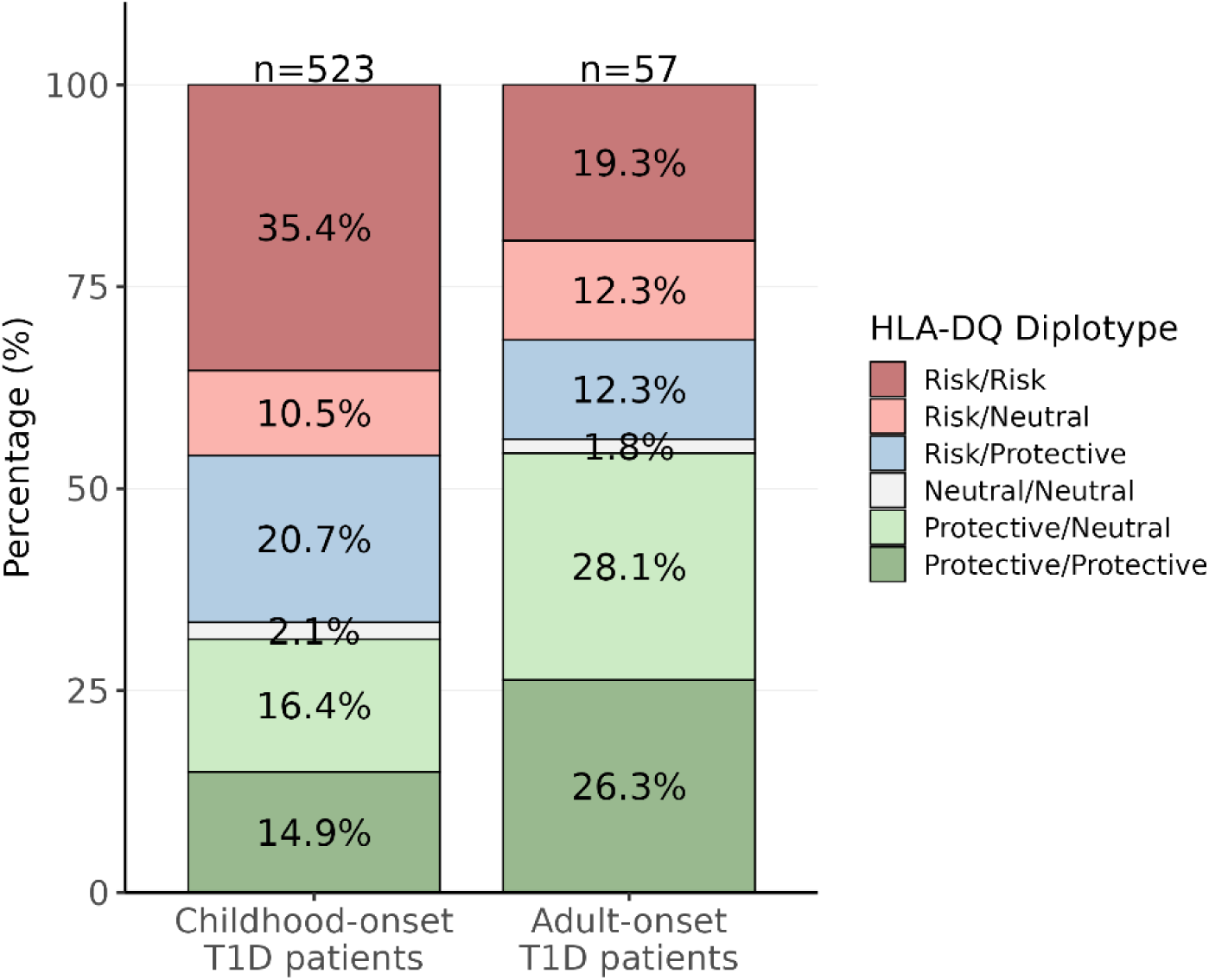
Stacked bar plot of *HLA-DQ* diplotypes stratified based on the risk, neutral and protective diplotypes. T1D, Type 1 diabetes. *HLA,* Human leucocyte antigen.

**Supplementary Figure 7:**
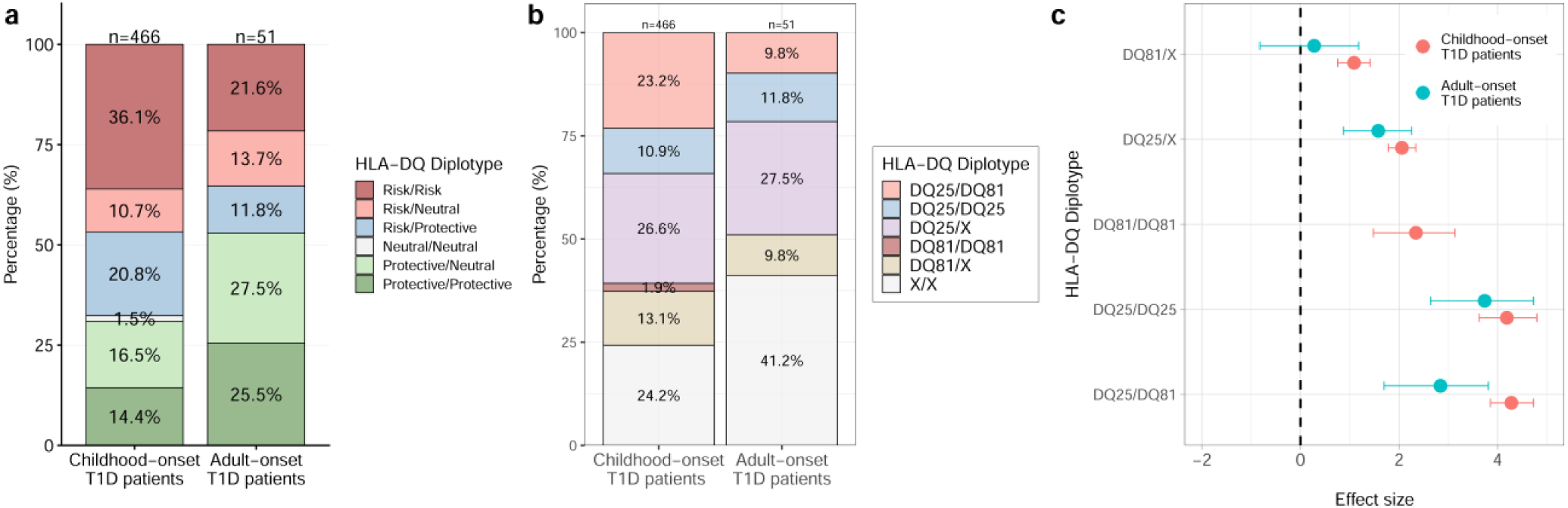
Comparison of HLA DQ diplotypes frequency and association analysis using a strict definition of T1D patients. **(a)** Stacked bar plot of HLA-DQ diplotypes stratified based on the risk, neutral and protective diplotypes. **(b)** Stacked bar plot of HLA-DQ diplotypes stratified by childhood and adult-onset T1D. **(c**) Forest plot showing the association status of T1D-associated HLA-DQ risk diplotypes. X represents any haplotype other than DQ25 or DQ81. T1D, Type 1 diabetes. HLA, Human leucocyte antigen.

**Supplementary Figure 8:**
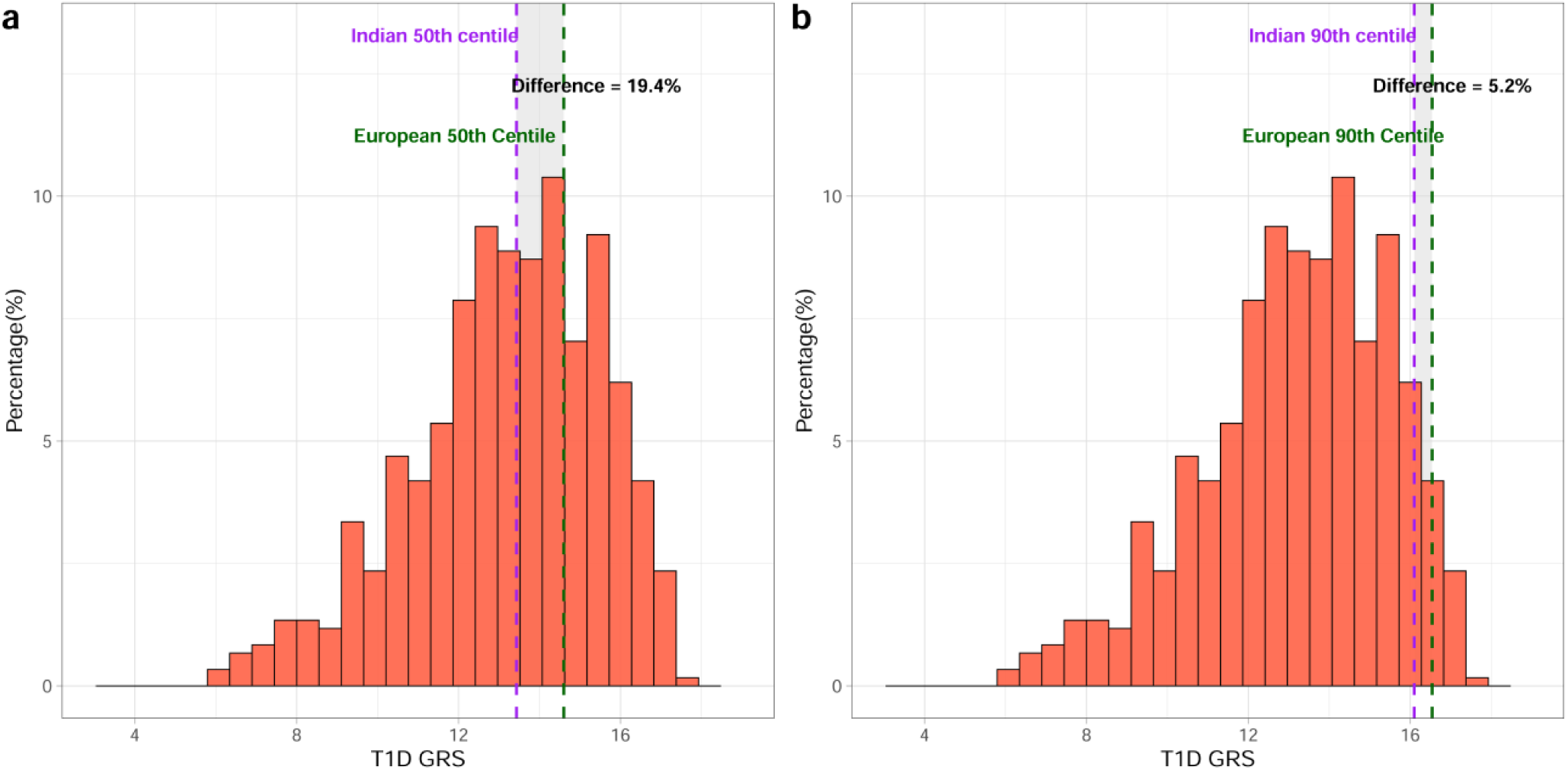
Comparison of Indian and European centiles of T1D GRS in Indian T1D patients. **(a)** 50^th^ centile of T1D GRS **(b)** 90^th^ centile of T1D GRS. T1D, Type 1 diabetes. GRS, Genetic risk score.

## Supplementary Tables

**Supplementary Table 1:** Clinical characteristics of the cohorts used in the study.

| **Group** | **T1D-CDI** | **T1D-TDRC** | **T1D-JSS** | **T1D-OSM** | **MMNP** | **MPC** |
| --- | --- | --- | --- | --- | --- | --- |
| **N** | 101 | 129 | 162 | 205 | 2300 | 1045 |
| **Males (%)** | 51.5% | 38.8% | 36.4% | 42.0% | 46.7% | 49.0% |
| **Age at Diagnosis\***<br>**(Yrs)** | 14.0<br>(11.0 - 21.1) | 10.0<br>(6.0 - 13.0) | 8.0<br>(4.0 - 11.0) | 9.0<br>(6.0 - 12.0) | NA | NA |
| **Age at Collection**<br>**(Yrs)** | 24.0<br>(14.8 - 29.2) | 17.0<br>(12.0 - 21.0) | 12.5<br>(9.0 - 15.0) | 13.0<br>(10.0 - 18.0) | 35.3<br>(30.4 – 39.8) | 32.0<br>(28.0 - 37.0) |
| **Duration of**<br>**Diabetes** | 6.0<br>(2.0 - 12.0) | 8.0<br>(4.0-11.0) | 4.00<br>(1.0 - 6.1) | 3.0<br>(1.0 - 4.5) | NA | NA |
| **C-peptide<sup>§</sup>**<br>**(pmol/L)** | 26.4<br>(3.3 - 141.9) | ND | 28.1<br>(6.6 - 159.0) | 26.48<br>(23.17 -29.79) | NA | NA |
| **GAD positive(%)** | 77.3% | 50.0% | 68.0% | 92.0% | NA | NA |
| **At least one islet**<br>**cell autoantibody**<br>**positive (%)** | 87.1% | 58.3% | 76.0% | ND | NA | NA |

Data shown as Median (IQR) unless indicated. NA represents not applicable. ND indicates not done. \*Data missing on 17 individuals <sup>§</sup>Data is missing on 5 individuals. T1D, type 1 diabetes. CDI, Chellaram Diabetes Institute, TDRC, Tapadia Diagnostic Research Centre. JSS, JSS Academy of Higher Education and Research. OSM, Osmania General Hospital. MMNP, Mumbai Maternal Nutrition Project. MPC, Mysore Parthenon Cohort.

**Supplementary Table 2:**
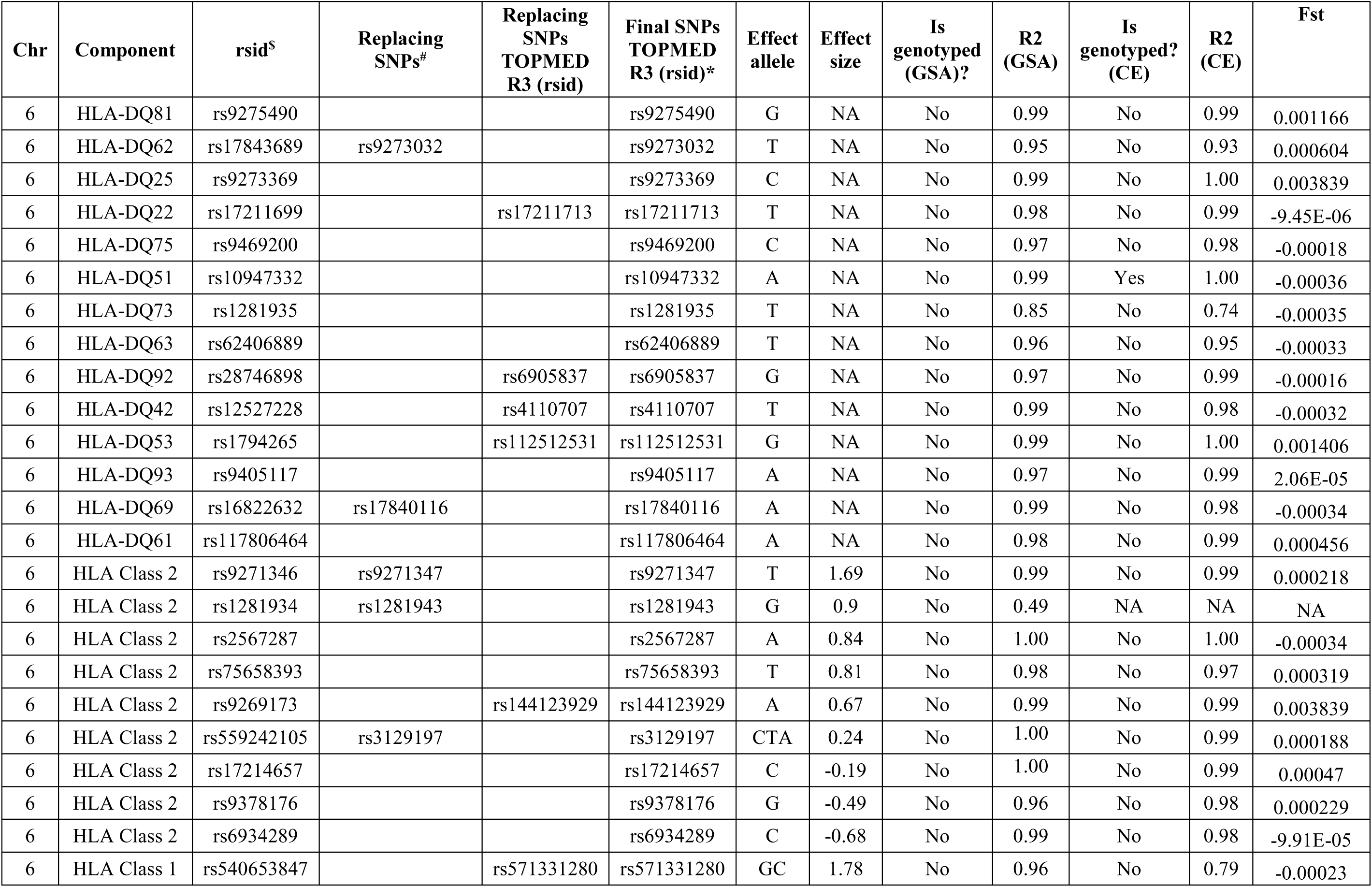
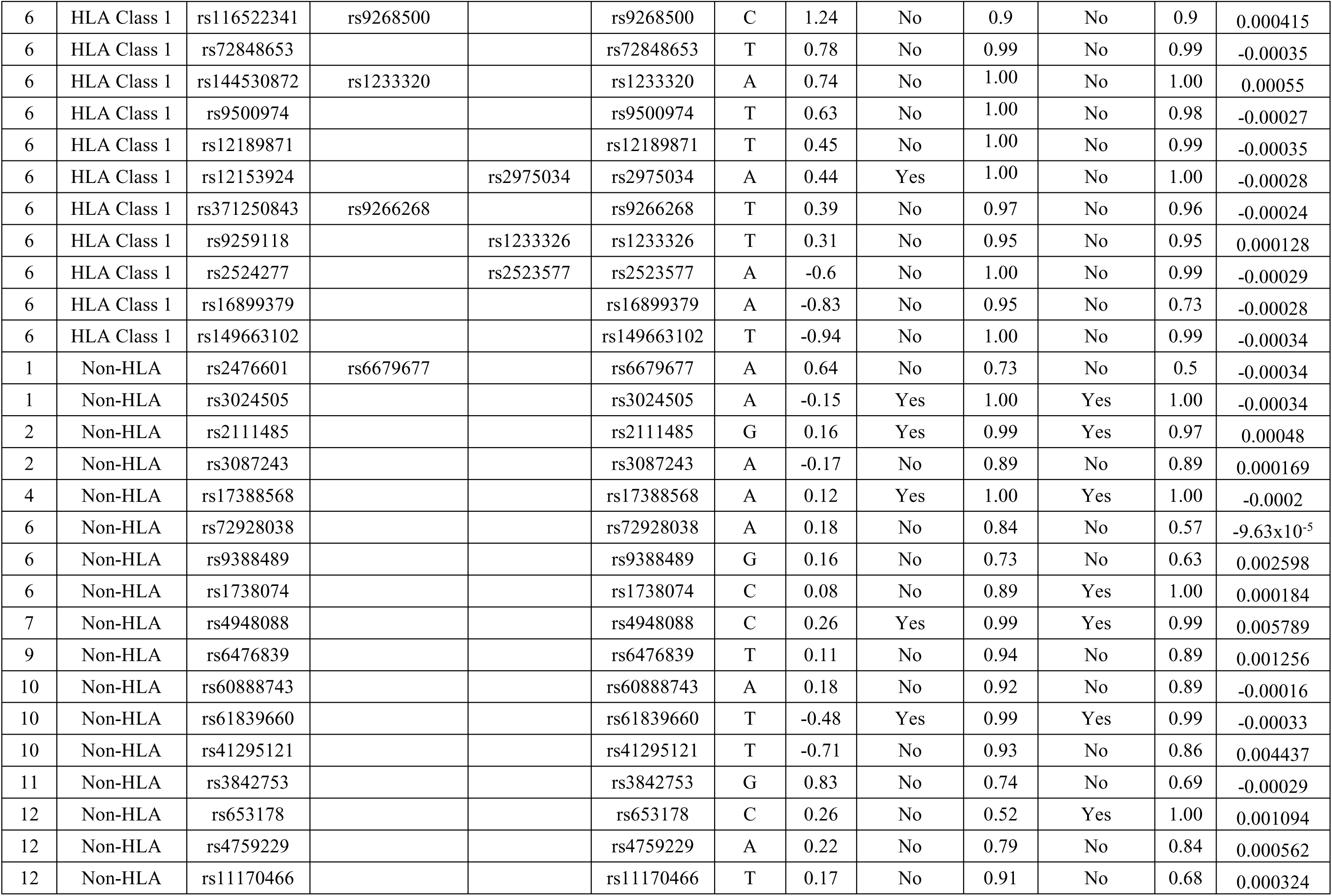
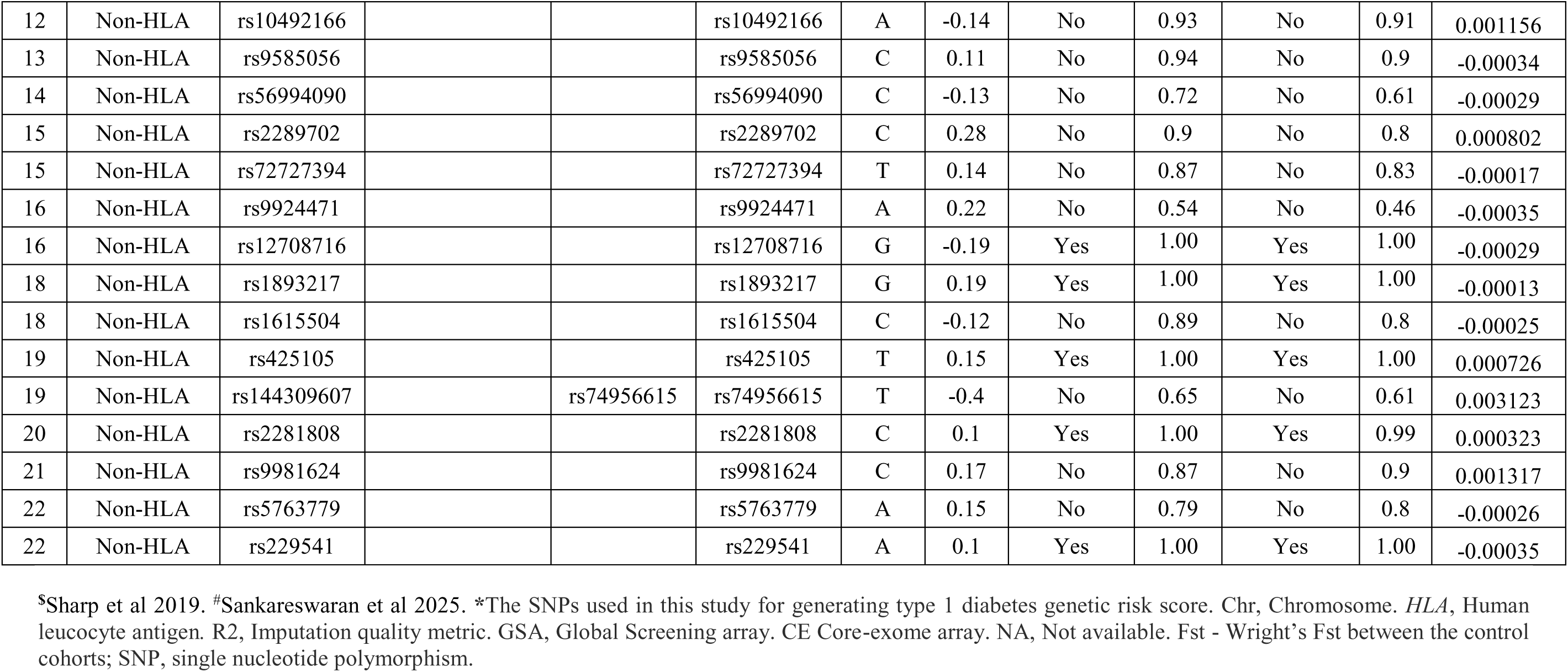
Details of SNPs used in type 1 diabetes genetic risk score.

**Supplementary Table 3:** Discriminative ability of T1D GRS in the multi-centric data stratified based on islet autoantibody status. Missing data for 17 samples is excluded.

| GRS type | Islet cell auto-antibody status | n | AUC (95% CI) |
| --- | --- | --- | --- |
| T1D GRS | Positive | 468 | 0.86 (0.84 - 0.88) |
|  | Negative | 112 | 0.75 (0.70 - 0.80) |
| HLA GRS | Positive | 468 | 0.85 (0.83 - 0.87) |
|  | Negative | 112 | 0.74 (0.69 - 0.79) |
| Non-HLA GRS | Positive | 468 | 0.61 (0.59 - 0.64) |
|  | Negative | 112 | 0.59 (0.54 - 0.64) |

T1D, Type 1 diabetes. *HLA*, Human leucocyte antigen.

**Supplementary Table 4:** Distribution of *HLA-DQ* haplotypes in T1D patients and controls, and their imputation accuracy.

| **DQA1~DQB1**<br>**haplotype** | **Controls (n (%))** | **T1D patients (n (%))** | **Avg. PP** |
| --- | --- | --- | --- |
| 01:01~05:01 | 192 (5.7%) | 30 (5.0%) | 0.90 |
| 01:02~06:02 | 75 (2.2%) | 3 (0.5%) | 0.83 |
| 01:02~06:09 | 57 (1.7%) | 5 (0.8%) | 0.84 |
| 01:03~06:01 | 590 (17.6%) | 22 (3.7%) | 0.83 |
| 01:03~06:03 | 159 (4.8%) | 5 (0.8%) | 0.85 |
| 01:04~05:03 | 389 (11.6%) | 67 (11.2%) | 0.80 |
| 02:01~02:02 | 471 (14.1%) | 40 (6.7%) | 0.89 |
| 02:01~03:03 | 160 (4.8%) | 3 (0.5%) | 0.89 |
| 03:01~03:02 | 347 (10.4%) | 142 (23.8%) | 0.87 |
| 04:01~04:02 | 18 (0.5%) | 1 (0.2%) | 0.87 |
| 05:01~02:01 | 214 (6.4%) | 240 (40.2%) | 0.89 |
| 05:05~03:01 | 132 (3.9%) | 7 (1.2%) | 0.83 |
| Others | 541 (16.2 %) | 32 (5.4%) | 0.83 |

Avg. PP represents the average posterior probability of HLA imputation. T1D, Type 1 diabetes. *HLA*, Human leucocyte antigen.

**Supplementary Table 5:** Counts of the *HLA-DQ* diplotypes.

| ***HLA-DQ***<br>**diplotypes** | **Adult-onset**<br>**T1D**<br>**patients (n)** | **Childhood-**<br>**onset T1D**<br>**patients (n)** | **Control**<br>**(n)** |
| --- | --- | --- | --- |
| DQ25/DQ81 | 5 | 117 | 33 |
| DQ25/DQ25 | 6 | 57 | 17 |
| DQ81/DQ81 | 0 | 11 | 19 |
| DQ25/X | 15 | 137 | 345 |
| DQ81/X | 5 | 69 | 448 |
| X/X | 26 | 132 | 2475 |

T1D, Type 1 diabetes. *HLA*, Human leucocyte antigen.

**Supplementary Table 6:** Comparison of HLA-DQ risk diplotype associations in T1D patients.

| *HLA-DQ*<br>diplotypes | Odds<br>Ratio | 95% CI | P | T1D<br>group |
| --- | --- | --- | --- | --- |
| DQ25/DQ81 | 72.24 | 47.47 - 113.30 | $4.10 \times 10^{-83}$ | Childhood-onset T1D |
| DQ25/DQ25 | 66.02 | 37.71 - 121.51 | $3.46 \times 10^{-45}$ | |
| DQ81/DQ81 | 10.38 | 4.39 - 23.10 | $1.93 \times 10^{-8}$ | |
| DQ25/X | 7.85 | 5.93 - 10.38 | $2.20 \times 10^{-47}$ | |
| DQ81/X | 2.97 | 2.14 - 4.10 | $7.55 \times 10^{-11}$ | |
| DQ25/DQ81 | 17.12 | 5.47 - 45.15 | $7.93 \times 10^{-8}$ | Adult-onset T1D |
| DQ25/DQ25 | 42.10 | 14.01 - 113.30 | $1.05 \times 10^{-12}$ | |
| DQ25/X | 4.85 | 2.39 - 9.58 | $6.49 \times 10^{-6}$ | |
| DQ81/X | 1.32 | 0.44 - 3.25 | 0.58 |  |

T1D, Type 1 diabetes. CI, Confidence intervals. *HLA*, Human leucocyte antigen.

**Supplementary Table 7:** Threshold estimations of T1D GRS based on T1D centiles.

| **T1D group centile** | **T1D GRS cutoff** | **Control Centile** | **Sensitivity** | **Specificity** | **Youden's Index** |
| --- | --- | --- | --- | --- | --- |
| 5 | 8.91 | 31 | 0.95 | 0.31 | 0.26 |
| 10 | 9.93 | 50 | 0.90 | 0.50 | 0.39 |
| 25 | 11.85 | 79 | 0.75 | 0.79 | 0.54 |
| 50 | 13.43 | 94 | 0.49 | 0.94 | 0.43 |
| 75 | 15.14 | 99 | 0.23 | 0.99 | 0.22 |
| 90 | 16.10 | 100 | 0.09 | 1.00 | 0.08 |
| 95 | 16.43 | 100 | 0.05 | 1.00 | 0.04 |

T1D, Type 1 diabetes. GRS, Genetic risk score.

